# Enhancing household Soybean Processing and Utilization in the Eastern Province of Zambia, a concurrent triangulation study design

**DOI:** 10.1101/2023.02.23.23286345

**Authors:** Funduluka Priscilla, Hachibamba Twambo, Mukuma Mercy, Bwembya Phoebe, Keth Regina, Kumwenda Chiza, Mwila Natasha

**Author notes:** Conceptualization, Writing – original draft, Visualization and Methodology, Research administration, Data collection, Analysis, Investigation, Software and Supervision. Reviewing and editing. Conceptualization, Visualization and reviewing Methodology.

## Abstract

Soybean processing and utilization is still low in the Eastern Province of Zambia despite the support by local and international NGOs in the sector. This study was designed to establish best ways of improving household soybean use in Petauke, Katete and Chipata districts in Eastern Province. In a concurrent triangulation study design, information was generated using a structured questionnaire on a sample of 1,237 households. Meanwhile two separate semi structured questionnaires were administered on a sample of 42 Focus Group Discussion and In-depth Interview (FGD and IDI) participants. Stata MP 15 (StataCorp, College Station, TX, USA) software generated frequencies and associated factors for soybean processing and utilization. NVIVO software QSR10 (QSRInt, Melbourne Australia) was used to organize qualitative data. All data was ultimately analyzed using the three Food Systems Thematic Areas namely; Socioeconomic, Enabling and Food environment as well as general environmental factors. Among the soybean products, a readymade Textured Vegetable Protein was universally consumed [1,030/1237(83%)]. Milled Soybean used for cooking Porridge and Shim [279/1237(22.55%)] among the few whole soybean products, emerged congruent with common household meals. However, accessibility of soybean for household consumption throughout the year was negligible. Immediate strategies should include; intensifying soybean production without leaving behind female headed households. Wards with poor markets, poor soils, and lack of land to grow could be market avenues for locally processed soybean products. Male involvement should be re-examined to improve on Soybean accessibility for household consumption. Educating farmers on the benefits of soybeans as well as how to make various soybean products should be strengthened. Dangers of anti-nutritional factors and how to destroy them in soybeans to ensure protein digestibility as well as promoting use of whole as opposed to refined mealie meal to ensure protein complementarity should be among the key messages. Market linkages of farmers to seed companies need to be created and strengthened with farmers themselves becoming more involved in soybean seed production and multiplication in order for them to access inputs at fair price. More companies should be encouraged to venture into soybean value addition, including privately owned community equipment such as hammer-mills. Extension services for soybean processing and utilization should be improved through training the camp officers, rural health facility outreach staff as well as community volunteers correct processing of various soybean products. Likewise, there is need to advocate among stake holders for more emphasis on soybeans in the food system. In the medium and long run, other equipment that could be promoted in the community include; Soybean oil expeller or press and Soybean blenders or Pulverizing equipment for making Soy milk.

## INTRODUCTION

Soybean processing and utilization is still low in the Eastern Province of Zambia. This is despite the support received from the Green Innovation Centers (GIC) under Deutsche Gesellschaft für Internationale Zusammenarbeit (GIZ), Food and Nutrition Security Enhanced Resilience (FANSER) Projects and other international and local projects, in the Soybean value chain. Soybean as a food crop provides several therapeutic benefits as Soy protein contains most of the essential amino acids in the amounts needed for human health [1]. These are histidine, isoleucine, leucine, lysine, methionine, phenylalanine, threonine, tryptophan and valine [1]. This is the reason why it is often called the “golden miracle bean,”[2]. Soybean is a leguminous crop because of its ability to form nodules and fix nitrogen in the soil just like any other legume [3]. It is botanically known as Glycine max and is a Climate-resilient low-cost crop increasing its potential as a food security crop [4], [5]. Soybean production occurs at a substantial scale in Zambia being the second-largest soybean producer in Southern Africa illustrating the important economic role that the crop plays in the country [6]. Eastern province although comprised mainly of small-scale farmers, is one of the three main producers of soya beans in Zambia. Others are Central and Northern Provinces [7], [8]. Soybean has unique characteristics as it can be made into a variety of products and processing it can be a means of income generation for households as well as ensure food security [9].

Popular household and industrial Soybean products include Soy yogurt, Soy milk and Soy cheese [1], [9], [10]. Soy flour, weaning food formulations, Soy based soups, confectioneries, beverages, fermented soy products and extruded products have also been documented [11]. These products are also commonly used in Zambia. Other known products are Soy relishes. Soy coffee, Soy sausage and Soy sprouts as well as tempeh, Soy sauce, Soy candies and Soy meat [9], [12]. Fermented products from soybean form a significant portion of the diets of the Asian populations and the consumer acceptability of such products is specific to countries or regions on account of their characteristic flavor [13]. “Tofu,” “miso,” “natto,” and “kinako” are popular Soybean foods in Japan, whereas tempeh is popular in Indonesia and China [13]. In Zambia Soybean is generally added to family meals, cooking oil and feed [6]. However household Soybean processing and utilization is still low in the Eastern Province of Zambia despite the province being among the three leading soybean producers [8]. The region also receives support from the Green Innovation Centers (GIC) under Deutsche Gesellschaft für Internationale Zusammenarbeit (GIZ), Food and Nutrition Security Enhanced Resilience (FANSER) Projects and other international and local projects, in the Soybean value chain.

A number of factors associated by Soybean processing and utilization have been documented. These can be categorized as socioeconomic, enabling environment, food environment and general environmental factors based on the food systems [5]. Food systems embrace the entire range of actors and their interlinked value-adding activities involved in the production, aggregation, processing, distribution, consumption, and disposal (loss or waste) of food products that originate from agriculture (incl. livestock), forestry, fisheries, and food industries, and the broader economic, societal, and natural environments in which they are embedded [14]. Socioeconomic factors include; awareness, production costs, access to market, diversification at the farm and membership to farmer’s organization as well as sociodemographic factors and technology. Awareness factors include; lack of awareness regarding health benefits of the Soybean products and effective ongoing training on how to process Soybeans as well as level of education [5], [12], [15], [16]. Production factors are farm size, cost of improved soya bean seeds, low fertilizer use as well as affordable credit services were also reported [7], [17]–[20]. Market related factors are farmers’ inability to access favorable Soybean outputs and processing markets [7], [15], [17], [21]. Farm diversification such as off-farm income, ownership of livestock as well as membership to farmer’s organizations [16]–[20]. Sociodemographic factors include age, occupation and gender. Furthermore limited soybean farm mechanization and soybean processing technologies [15].

Environmental factors influencing Soybean Processing and Utilization are three fold. These are enabling environment, food environment and general environment. Enabling environment factors include; yield of Soybean, access to technologies for production, harvesting and processing [5], [9], [11], [17]. Others are access to agricultural advisory services, household size as well as value chain governance [7], [9], [12], [17], [19], [20] Food environmental factors include; indigestion problems, characteristic beany, presence of anti-nutritional factors as well as the hard-to-cook characteristics associated with Soybean [9], [15], [17]–[20], [22]. General environment factors include; poor soils in some cases and Soybean cultivation being mostly rain-fed [7], [15]. Overall, integrated programs involving all stakeholders in the food system are recommended [5].

The purpose of this study was to determine the best ways of improving household soybean processing and use among households in Eastern province for adoption by the GIZ funded projects as well as other projects operating in similar environments. To the best of our knowledge this is the first research in this regard in Zambia.

## METHODOLOGY

### Study Setting

This study was conducted in the Eastern Province one of Zambia’s ten provinces [23]. The province lies between Malawi to the east and Mozambique to the south [23]. Locally it shares borders with three other provinces of the country, namely, Lusaka, Central and Muchinga Provinces [23]. With the provincial capital being Chipata, eastern province has an area of 51,476 km2. A population of 1,592,661, accounting to 12.16% of the total Zambian population was recorded in 2010 with 1,030 for females every 1,000 males [24]. Chewa was the largest community in the region and the most widely spoken language with 34.6 per cent people speaking it [24]. The study was conducted in the three districts. These are Petauke, Katete and Chipata. The predominant economic activity is farming [24].

### Study design

In this study all data were collected at the same time and later triangulated thereby generating rich information to inform programming, thereby making the concurrent triangulation study design appropriate [25]. Two other study designs were nested in this strategy. A cross sectional survey was used to collect quantitative information on factors associated with Soya bean processing and utilization; and a barrier analysis was used to generate in-depth data that provided additional insight.

### Cross sectional survey

A cross-sectional survey generated information on factors associated with Soybean processing and utilization using a structured questionnaire. The questionnaire captured information on Socio-demographic characteristics as well as associated factors for Soybean processing and utilization based on the food systems. A pilot study was conducted in Petauke in order to ensure accuracy and reproducibility of the results. Up to 168 households that never participated later in the study were enrolled. This was followed by questionnaire update in the kobo collect toolbox, which was redeployed in real time for accessibility and data collection immediately using smart phones. The questionnaire was finally administered completely on 1,237 households out of 1258 households planned for in the study giving a non-response rate of 1.67%. This rate is lower than that recorded (4%) in one the national household surveys [26].

Sampling for the Cross Sectional Survey

Here the study adopted the Yamane (1967) technique, which states that the sample size n is defined as:

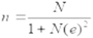

Where n is the sample size, N is the population size and e is the level of precision. At 95% confidence level, e = 0.05. The sample size was adjusted for non-response rate. The prediction of non-response rate was based on Zambia Demographic Health Survey non response rate [26]. Each district had a specific sample size calculated based on their population size. The population figures for calculating sample sizes were obtained from the city population website for 2016. Yamane’s technique was appropriate in this case as there was no known prevalence of Soybean utilization.

### Selecting clusters for cross sectional survey

In this study, five clusters from each district were included in the study. A cluster was defined as a ward, which is an official administrative unit under a district in Zambia. The number of wards selected was matching with five working days in each district. The wards were segmented using camps as boundaries manned by camp officers under the Ministry of Agriculture. This was in order to effectively collect the required data. The five clusters for each district were randomly assigned by probability proportional to size (PPS) using the ENA software [27]. The selection of the clusters was conducted at district level after excluding the wards that were based in town. One segment from each ward was selected according to PPS. In each segment, PPS was also used to select villages. In cases where the targeted number of households was not reached, the adjacent village was included in the study.

### Selecting Households for Cross Sectional Survey

This study was conducted at the beginning of the planting season and most households went out every morning to prepare fields and gardens. The preferred sampling method in this case therefore was convenient sampling. This meant that as soon as a household was available, they were recruited and were interviewed. Informed consent was obtained from all before commencement of the interview. The participants were also informed that they were free to decide not to take part in the interview and also to with draw at any time. There were no risks for participants in this study. Participants were invited to participate as the findings were necessary to shape the best ways of processing and utilizing soybeans in households to improve on dietary diversity and ultimately nutrition status. These ethical considerations are according to the Helsinki declaration of 1964 on studies involving human subjects [28]. Some of the respondents were followed in their fields during demonstrations organized by agricultural extension officers.

### Training Data Collectors for Cross Sectional Survey

A one day training workshop for survey team members was conducted. The training covered general survey objectives, overview of survey design, household selection procedures, data collection and interview skills. The objective was being able to ask the questions in the questionnaire correctly. The survey team members were closely monitored during data collection in the field

### Data analysis for Cross Sectional Survey

Quantitative data was downloaded from the kobo collect toolbox in the format suitable for excel sheet (XLS legacy). This was then cleaned and imported into the Stata software coded and analyzed. Frequencies were used to describe some socioeconomic and sociodemographic characteristics of participants, household soybean production, processing of whole soybean and utilization of whole and ready-made soybean products. Mean (SD) was used to describe age. All variables which included; Socio-demographics, socioeconomic, knowledge attitudes and practices were later fitted into the multiple logistic regression model to come up with the adjusted estimates in the most efficient model that rules out confounding factors. The variables both significant and those not significant at <0.05 were entered using weighted logistic regression. After controlling for all the other factors a number of them were found to be associated with soybean processing and utilization (p<0.05). All the factors associated with soybean processing and utilization were entered into the best fit (final) model in order to report Adjusted Odds Ratio with 95% Confidence Interval.

### Barrier Analysis

This is a Participatory Analysis Tool that identifies key enablers and barriers to the implementation of practices in resource-poor communities [29]. In order to generate information on the barriers or enablers for soya bean processing as well as utilization, Focus Group Discussions and In-depth Interviews (FGDs and IDIs) functioned as data collection instruments with the help of semi structured question guides. Focus group discussions were chosen in order to reveal collectively shaped social processes [30]. On the other hand In-depth Interviews were chosen to get a vital source of information being the perspectives of individuals who have personal experiences with Soybean processing and utilization [31]. Barrier Analysis was conducted by going through the procedure in **Table 1**.

**Table 1.**
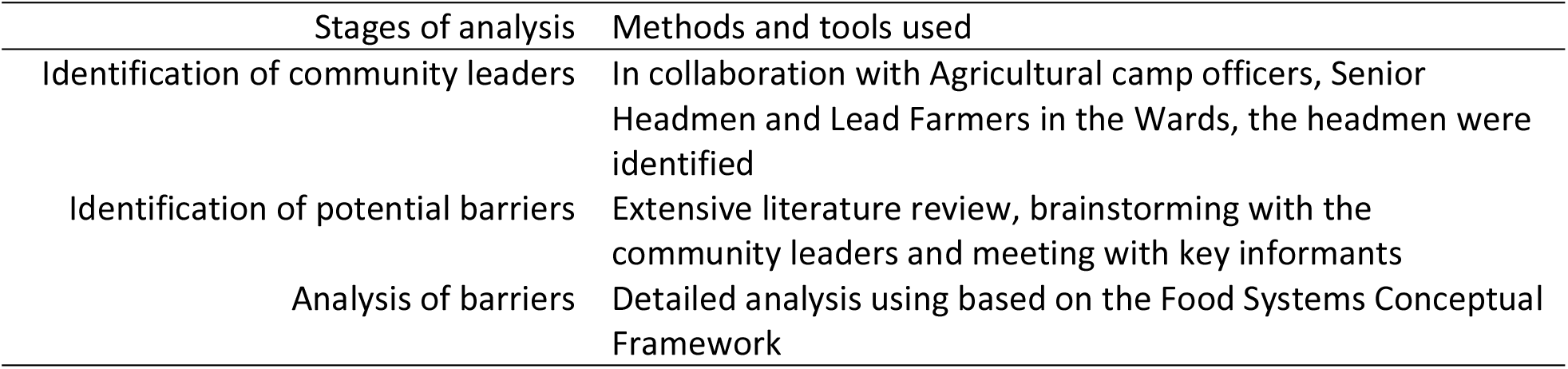
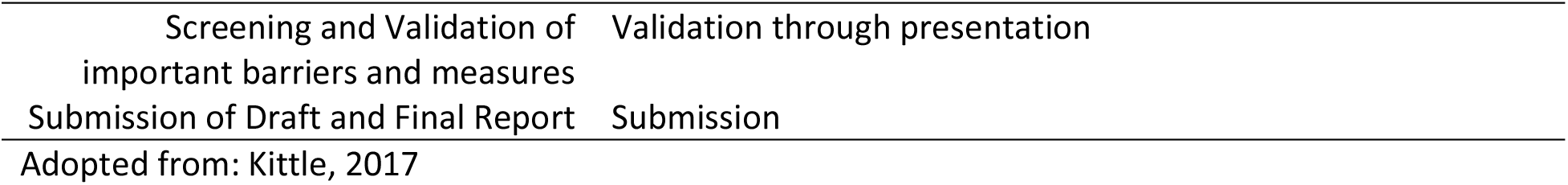
Conducting Barrier Analysis

### Sampling for Barrier Analysis

Community leaders were purposively sampled for Focus Group Discussions (FGDs). The number of focus group discussion (FGDs) were determined by theoretical saturation [32]. This means that participants for FGDs were recruited and Discussions were held until a point where there was no more additional information being generated [32]. In-depth interviews (IDIs) were equally conducted. IDIs are a tool for collecting rich information that can inform program development and evaluation [33]. Key Informants were purposively sampled. These came from the Ministry of Health and Ministry of Agriculture Officials as well as GIZ. Informed Consent was obtained from all participants before interviews.

Each FGD was conducted by two moderators and digitally recorded. Permission to record the discussions digitally was sought from all participants. The discussion were moderated by one facilitator who also ensured that all the topics were covered in the interview guide. A note-taker assisted with recording both digitally and by writing which helped in determining emerging themes. Each FGD lasted for an average of an hour. FGD venues used were mainly meeting sites for farmers with Camp officers, which are open places away from houses. Two out of six FGDs were conducted in a room at a health facility as well as in a classroom at a primary school. All IDIs were conducted via telephone. This is because it was not possible to meet participants physically due to distances as well as busy work schedules.

### Data Appraisal for Barrier Analysis

Barrier analysis results were categorized based on the food systems thematic areas. These are social economic factors, factors in the enabling and food environment as well as general environmental factors. This was carried out using NVIVO QSR10 (QSRInt, Melbourne Australia). Reading transcripts repeatedly ensured gaining a deeper insight of the data [34]. Coding was the next stage. A code is a word or sentence or phrase that represents aspects of a data or captures the essence or features of a data [35]. Codes were then matched with segments of text or informant statements selected as representative of the code as recommended [36]. The original meaning of what was communicated by the informants was maintained.

### Ethical Considerations

This research was approved by the Levy Mwanawasa Medical University Research Ethics Committee (LMMU-REC 00010/20) as well as the National Health Research Authority (NHRA). Permission was obtained from Petauke, Katete, Chipata, Chipangali as well as Kasenengwa district agricultural coordinators’ offices. Permission to collect data was also obtained from the senior headmen in each ward. Since this was a low risk research informed consent was verbally obtained from respondents. Respondents were selected and interviewed in their homestead away from other family members in order to ensure privacy. The respondents were free to withdraw from the study at any time. Data collected was de-Identified.

## RESULTS AND DISCUSSION

### Characteristics of participants

#### Socio-demographics of Cross Sectional Survey Respondents

Up to 1,237 out of 1258 households participated in the study giving a non-response rate of 1.67%. The mean age of respondents was 40.33(SD=13.63) years. Male headed households were 780/1237(63%) being more than female headed households which were 457/1237(37%). National values for male headed households are at 74.2% slightly more than what was reported in this study. This also shows that male headed households are more in Zambia compared to female headed households which are at 25.8% (CSO, 2018). Eastern Province the study site is also known to have more females than males. This is confirmed by the sex ratio of 2010 census which was 1,030 for every 1,000 males (CSO, 2010). Despite all this up to 733/1,237(59.26%) females more than males [504/1,237(40.74%)] participated in the Cross Sectional Survey. This could mean that females were found in the households during the survey while the males were mostly absent. The majority [669/1,237(54.08%)] of the participants were Chewa speaking. These were found in substantial numbers in all the three districts and comprised almost the entire population [387/396(97.73%)] in Katete District. This is in agreement with the 2010 census whereby Chewa was found to be the largest community in Eastern Province with 39.7 per cent of the total population and Chewa was the most widely spoken language with 34.6 per cent speaking it (CSO, 2010). **Table 2** shows the details.

**Table 2.**
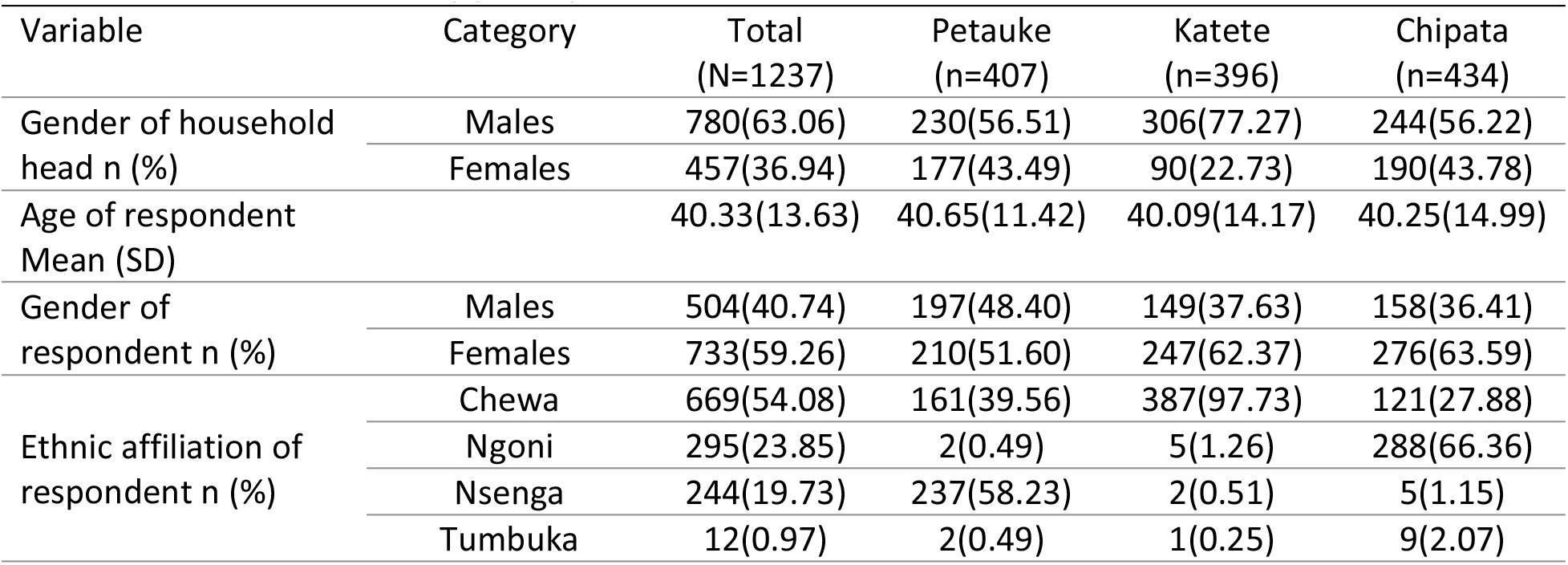
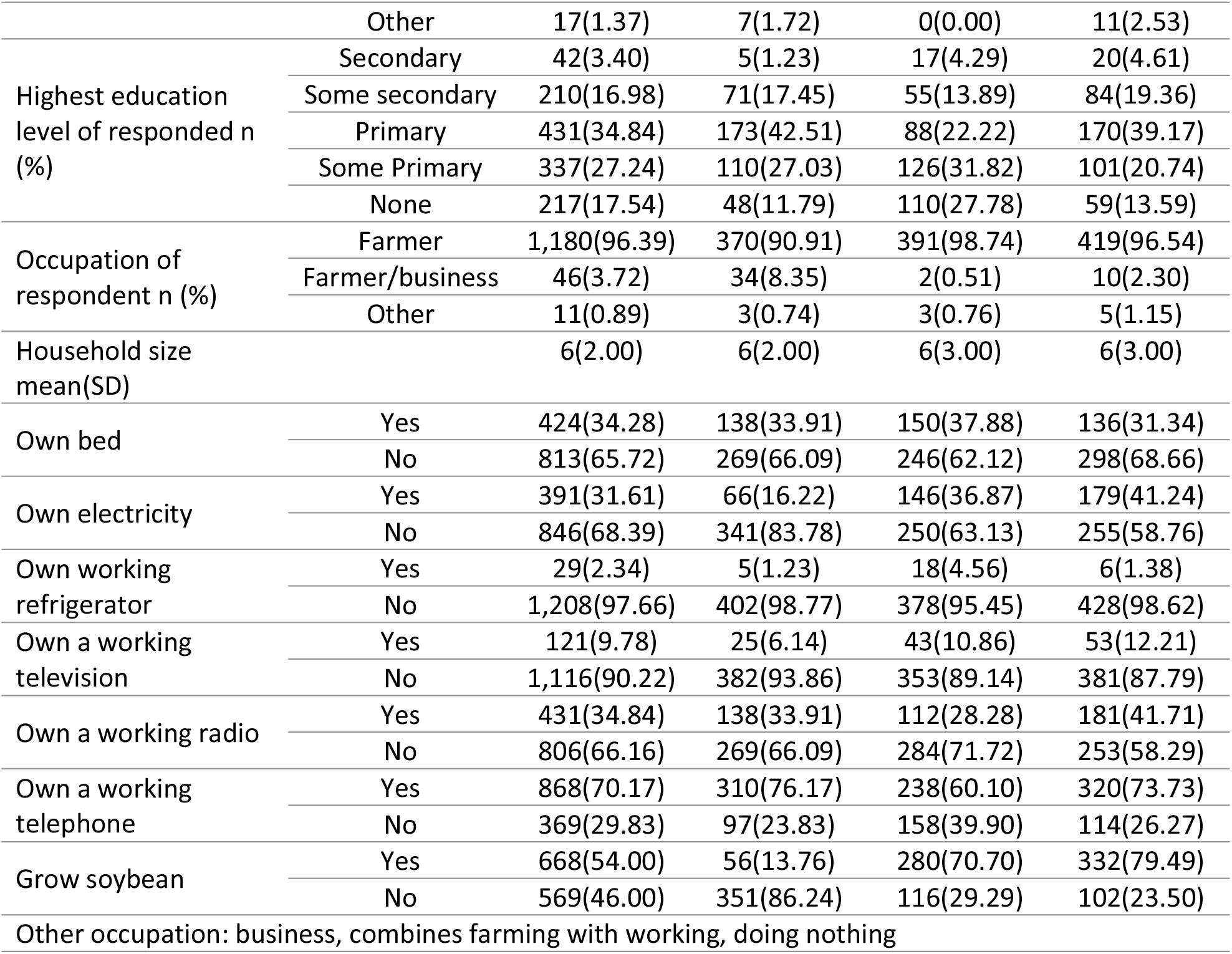
Cross sectional survey participants

#### Socioeconomic status of Cross Sectional Survey Respondents

The highest level of education reported by most respondents [431/1237(34.84%)] was primary education. Petauke with 173/1237(42.51%) had the highest proportion with Katete [88/1237(22.22%)] trailing behind. Katete also had a number of respondents with some primary as well as no education at all [126/396(31.82%)] and [110/396(27.78%)] respectively. This agrees with the Second Report of the Committee on Education, Science and Technology for the Fourth Session of the Tenth National Assembly appointed of 24 September 2009, which showed that among the districts with unacceptable levels of adult illiteracy, Katete was among the highest with 62.9%. Farming [1,180/1237(95.39%)] was the predominant occupation reported in the three districts. Some off farm business activities were documented in Petauke [34/407(8.35%)] and Chipata [10/434 (2.30%)]. Mean household size was 6(SD=2) higher than the national value of 5.2 reported for rural settings in the 2018 Zambia Demographic and Health Survey (CSO, 2018). Four major consumer goods owned in the three districts include a working mobile telephone, a working radio, owning a bed and having electricity (MTN set or solar panel) with 868/1237(70.17%), 431/1237(34.84%), 424/1237(34.28%), 391/1237(31.61) respectively (**Table 2**).

#### Socio-demographics of barrier analysis participants

There were six focus group discussions (FGDs). Two FGDs were conducted in each of the selected wards in the three study districts. Up to 36 participants took part in the FGDs. The median age of participants was 50(Range=28 - 75) years. The youngest participant as well as the oldest participant aged 28 and 75 years respectively were from Chipata district. In-depth Interview (IDI) participants were six. Their median age was 41(Range=36 - 57) years. **Table 3** highlights the details.

**Table 3.** Barrier analysis participants

| District | Planned number Participants | Actual Number of Participants | Gender |  | Ward/Organization | Data Collection Method | Age Range |
| --- | --- | --- | --- | --- | --- | --- | --- |
|  |  |  | Male | Female |  |  |  |
| Petauke | 20 | 16 | 14 | 2 | Chilimanyama Msumbazi | FGD | 32-72 |
|  | 2 | 2 | 2 | 0 | MoA COMACO | IDI | 36-57 |
| Katete | 20 | 8 | 7 | 1 | Katiula Chimtende | FGD | 31-67 |
| Chipata | 20 | 12 | 11 | 1 | Nthope<br>Makangila | FGD | 28-75 |
|  | 4 | 4 | 2 | 2 | MoA, FANSER,<br>GIZ, GIC | IDI | 38-47 |

MoA: Ministry of Agriculture; COMACO: Community Markets for Conservation Ltd; FANSER: Food and Nutrition Security Enhanced Resilience; GIC: Green Innovation Centers; GIZ: Deutsche Gesellschaft für Internationale Zusammenarbeit; FGD: Focus Group Discussion; IDI: In-depth Interview

#### Soybean production, processing and utilization Production

The overall proportion of respondents who reported growing Soybean were 668/1237(54%). These were more in Chipata [332/434(79.49%)] followed by Katete [280/396(70.70%)]. In Petauke only 56/407(13.76%) respondents re-counted growing Soybean. Perceptions of headmen showed that soybean growing varied in the districts under study. It was ranked as number one in Nthope ward in Chipangali the new district, which fell under Chipata in this study. It was ranked second in Chimutende ward in Katete. Meanwhile, in Makangila ward in Chipata and Katiula ward in Katete it was ranked third. On the other hand, in Petauke district, Chilimanyama ward ranked it fourth while soybean growing was not reported in Msumbazi ward also in Petauke. These findings are consistent with a report by Lubungu and colleagues which showed that one of the districts where soya bean production is concentrated in eastern province is Chipata with erratic participation reported in Katete and some other districts [8]. This study has also shown that soya beans growing reports were massive among the male headed households [466/668 (70%)] than among the female headed households [202/668 (30%)]. The incidence of adoption among the female-led households is low possibly because they are constrained by lack of access to input, credit, and extension services [2] (**Table 4**)

**Table 4.**
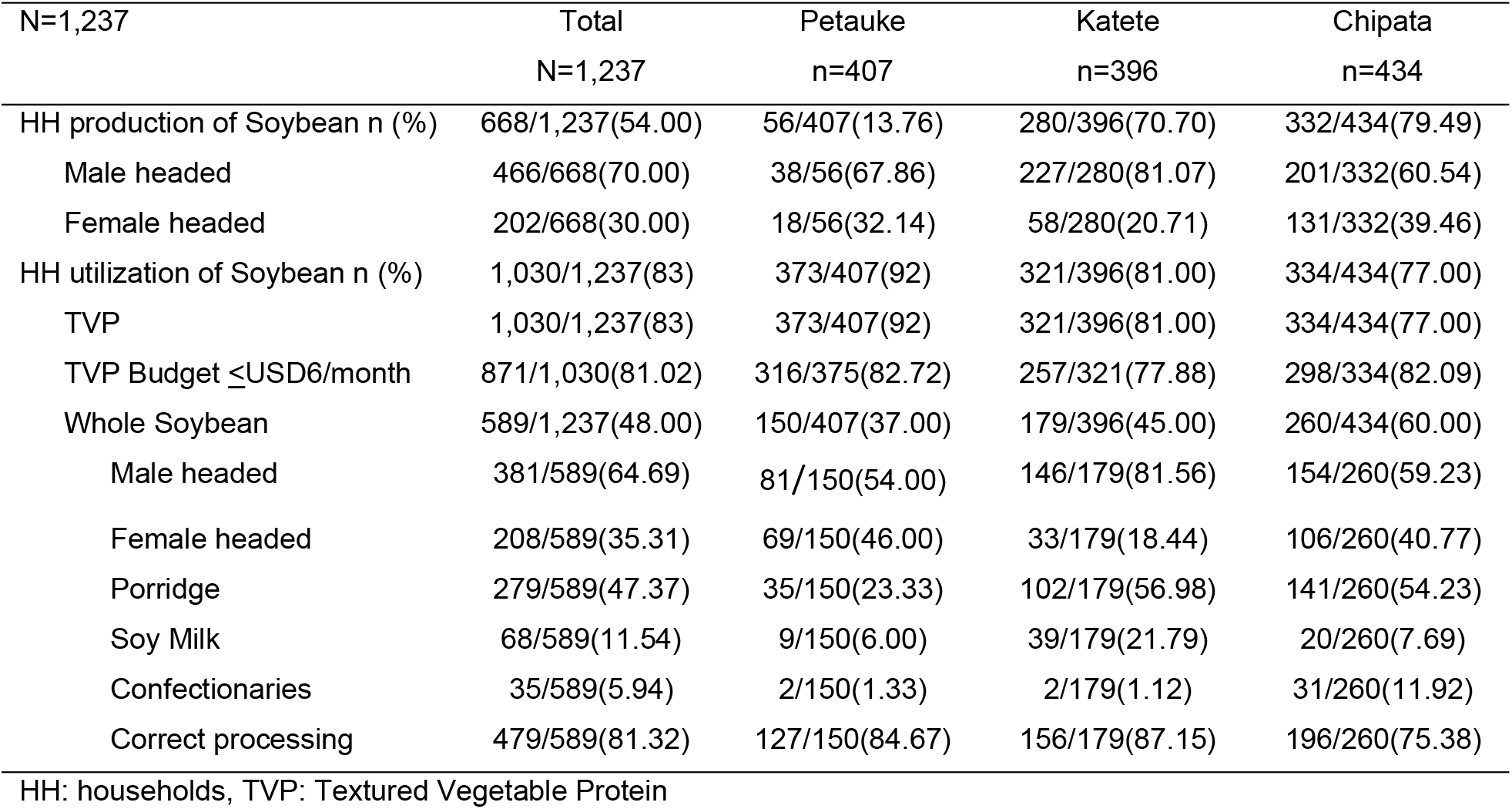
Household Soybean Production and Utilization

### Processing and Utilization

#### Utilization of ready-made soybean products

Textured vegetable protein (TVP) popularly known as Soya pieces or nyama soya in the study districts was the only ready-made product reported to be regularly consumed in the rural areas in the three districts. It was universally consumed [1,030/1237(83%)] as an easily accessible and relatively affordable relish to accompany a staple cereal thick porridge meal (known as nshima in Zambia). Varying consumption levels were reported. In Petauke reports were at 373/407(92%), with Katete and Chipata at 321/396 (81%) and 334/434(77%) respectively (**Table 4**). TVP was congruent with the dietary pattern in the study districts. The headmen reported that the common daily diet was nshima accompanied by a vegetable usually dry pumpkin leaves or okra, occasionally with textured vegetable protein based on the availability of money.

> “On a daily basis we eat nshima or samp, whole soybean is known for money we just leave seed, we only consume it in the form of soya pieces” (A Headman in Chimutende ward in Katete, FGD002)

In this study the least producer of soybeans, Petauke district is the highest consumer of the ready-made soybean product the TVP, with households spending up to USD15 per month. Households buy the product from the local retail outlets. This shows the willingness to buy one of the ready-made commercial soybean products. In addition it also shows that communities with disposable income such as those from Petauke with some of off farm activities as reported in this study are able to buy ready-made soybean products. It is also worth noting that most of the TVP found during the study period were from Malawi. The local shop owners however confirmed that they do occasionally stock one local brand. This could mean an unmet market gap by the local soybean value addition companies. Some whole soybean being exported to Malawi during the significant amount of trade in soybeans taking place across the Zambia-Malawi border [37] is processed there and comes back as TVP. One of the companies that locally make readymade soybean products is the Community Markets for Conservation (COMACO) a non-profit private company. The company is located in Chipata the provincial headquarters of Eastern Province with a branch in Petauke district. The products include yummy soy, crude oil and TVP. These are distributed in supermarkets and other retail outlets as well as to NGOs involved in school feeding programmes [8].

#### Processing and Utilization of whole soybean

In **Table 4** almost half [589/1237(48%)] of the respondents reported processing and utilizing whole soya beans in family meals. This proportion was arrived at by taking into account reports of making at least one whole soya bean product. Chipata ranked the highest with 260/434(60%), followed by Katete with 179/396(45%). Soya bean utilization was lower in Petauke being 150/407(37%). The differences in soya bean processing and utilization reports among the three districts were significant (p<0.0001) with Petauke explaining much of the difference. It should be noted here that the pattern of soya bean utilization among the three districts is similar to the pattern of soya bean growing. Soybean utilization is high where soya beans is produced more and therefore available, it is lowest where production is low. However, accessibility of whole soya beans for various products throughout the year was negligible [3/1030(0.29%)]. This means that the households were only able to make various products during the harvest season. Utilizing whole soya beans in family meals could bring about improvement in nutritional status in the households.

#### Whole soybean products at household level

The most frequently reported product prepared out of soybeans for family consumption was Soy Porridge. This was reported among 279/1237 (22.55%) of households. More reports came from Chipata [141/434(32.49%)] followed by Katete [103/396(26.01%)] and Petauke [35/407(8.60)]. The porridge was prepared out of the composite flour made by mixing maize with preheated or raw soybeans and then taken to the hammer mill. The milled flour was also sometimes used to make nshima and confectionary products to a limited extent. Confectionary products made from the composite flour of maize and soya bean producing cakes were known as ‘Vigumu’ in local language. Fritters were equally made from the soy flour mixed with wheat flour. These were reported to be sometimes sold within the neighborhood. Other products itemized at low levels include soya flour, boiled whole soybeans, soya sausage, and coffee and Soy milk (**Table 4**).

#### Correct whole soybean processing method reports

**Table 4** also shows households that reported correct soybean processing methods. Up to 479/724 (66%) reported having correctly processed whole soybeans by preheating it before mixing it with maize and taking it to the harmer-mill. These were more in Petauke being 127/134(95%) than in Chipata and Katete with 196/318 (62%) and 156/272(57%) respectively. Adverse nutritional and other effects following consumption of incorrectly processed or raw soybean meal have been attributed to the presence of endogenous inhibitors of digestive enzymes and lectins and to poor digestibility. To improve the nutritional quality of soy foods, inhibitors and lectins are generally inactivated by heat treatment [38]. Dry heat treatment can be applied at high temperatures above 120°C [39]. In a rural setting this could be achieved by roasting the dry cleaned soybean to slightly brown color.

#### Factors associated with soybeans processing and utilization at household level

**Table 5** shows a multivariate analysis. All the variables were first fitted in the multiple logistic regression model to come up with the adjusted estimates in the most efficient model that rules out confounding factors. The variables both significant and those not significant at <0.05 were entered using weighted logistic regression. After controlling for all the other factors a number of them were found to be associated with soybean processing and utilization (p<0.05). These are discussed in the sections that follow.

**Table 5.** Factors associated with whole soybean processing and utilization

| Variable | Multivariate Analysis |  |
| --- | --- | --- |
|  | AOR (95%CI) | P-Value |
| District | 0.56(0.41 0.75) | 0.00 |
| Male headed households | 1.88(1.15 0.06) | 0.011 |
| Male respondents | 0.48(0.30 0.76) | 0.002 |
| Being a Chewa | 1.17(1.08 1.26) | 0.000 |
| Owning a bed | 1.73(1.20 2.48) | 0.003 |
| Growing Soybean | 9.14(5.46 15.29) | 0.000 |
| Consuming Porridge/Nshima | 866.51(116.32 6454.96) | 0.007 |
| Processing barrier | 0.45(0.30 0.68) | 0.000 |

#### Factors that increase chances of soybean processing and utilization at household level

Male headed households AOR 2.15; 95% CI 1.21 to 3.79, being a Chewa AOR 1.26; 95%CI 1.15 to 1.38 and owning a bed AOR 1.78; 95%CI 1.16 to 2.74 were associated with increased chances of soybean utilization. In addition reporting preparing porridge or Nshima AOR 1905.14; 95%CI 241.50 to 15029.39, as well as Growing soybeans AOR 11.92; 95%CI 5.34 to 26.60 were equally associated with increased chances of soybean processing and utilization **(table 5)**. Among the households, male headed ones were reported to be growing more Soybean (70%) than those headed by females. Growing soybeans in this study was significantly associated with increased chances of utilization. In addition, Chewa speaking people had increased chances of processing and utilizing Soybean because they are the largest community in Eastern Province with 39.7 per cent of the total population as at 2010 population census. In Katete district they are almost the only ethnic group (97.73%). Katete district came out second in soya bean utilization in this study. Furthermore, Households who owned a bed had increased chances of processing and utilizing soybean as this is a sign of good social economic status in a rural setting. This could have been brought about by agricultural activities such as growing soybeans as a cash crop. In this study, 274//668(41%) of households that grew soybeans also owned a bed. Among the Soybean products, preparing porridge was one product that was significantly associated with Soybean processing and utilization AOR 866.51; 95%CI 116.32 to 6454.96. Porridge or Nshima are the Soybean products that most households (23%) attempted to make. In this case, whole soybeans is milled and mixed with maize and prepared as porridge or nshima. This could indicate that these products were compatible with meal patterns. FGDs with headmen confirmed this by showing that porridge and nshima were common staple meals thereby making it easy to adopt utilization of soybeans through these products. Once available, the composite flour (mixture of maize and soybean) was at times used to make soybean cake (Vigumu in local language).

#### Factors that reduce chances of soya bean processing and utilization at household level

District AOR 0.33; 95%CI 0.22 to 0.50, being a Male respondents AOR 0.41; 95%CI 0.24 to 0.72, as well as experiencing soybean processing barriers, AOR 0.42; 95%CI 0.26 to 0.69 reduced the chances of soybean processing and utilization **(table 5)**. Processing and utilization of whole soybeans was generally low in all the three districts. Petauke had the lowest prevalence being 150/589 (25%), followed by Katete with 179/589 (30%) and Chipata with 260/589 (44%). Meanwhile, only up to 249/1237 (20%) of the male respondents reported utilizing whole soybeans. It should also be noted that lack of knowledge was reported more among the male respondents as compared to females during the FGDs with the headmen. This means that most males in the three districts are ignorant about soybean processing into various products for family consumption. Low involvement of males in soybean utilization is consistent with existing literature [17]. They know it as a cash crop. This is despite the deliberate strategies in place especially in the GIZ implementation areas to include the men during household counselling in the CRS Care Group Model. There is need to re-examine the male involvement approach. A headman in Chimutende during FGD004 said “…men are not involved in soybean processing and utilization. If a man is not involved he will just want to sell”. On the other hand up to 670/1237 (54%) households experienced whole soybean utilization barriers. These are shown in **Table 5**.

#### Pooled barriers

Cross sectional survey as well as barrier analysis data have been pooled together using the food systems thematic areas to give the overall picture of barriers in soybean processing and utilization. Major barriers for Soybean processing and utilization were categorized as; Socioeconomic, enabling environment and food environment as well as general environmental factors. Socioeconomic barriers reported were lack of knowledge, Lack or unreliable soybean outputs and soybean processing markets, Lack of government control on soybean marketing, In addition, lack of equipment and ingredients, poor Soybean value chain governance and gender issues were also reported. Enabling and Food Environmental factors reported were; availability of Soybean for household consumption, accessibility of Soybean for household consumption as well as Safety, Health and convenience issues around Soybean utilization. While general environmental factors had to do with lack of land and poor soils (**Table 6**).

**Table 6.** Pooled Barriers for Soybean Processing and Utilization

| **Barrier Category** | **Broad/common barrier** | **Food system stage** |
| --- | --- | --- |
| **Socioeconomic** |  |  |
| **Knowledge** | Overall 39.55% of cross sectional respondents cited lack of knowledge as contributing factor for Soybean processing and utilization. | Soybean processing |
|  | Lack of awareness about health and nutritional benefits |  |
|  | Unstandardized soybean processing at household level |  |
|  | Inadequate coverage of existing projects & extension services on soya beans processing and utilization. Extension services as well as not being a member to existing programs |  |
| **Market led Processing and utilization for Soybean** | Lack or unreliable soybean outputs and soybean processing markets in some wards to enhance market led growth | Soya bean Production, Markets, Processing and household utilization |
| **Government Control** | Lack of government control on soybean marketing | Soybean Production, Markets, Processing and household utilization |
|  | Many textured vegetable protein products (TVP) come from Malawi. This is found in local shops and consumed a lot by local households | Soya bean Processing |
| **Equipment and ingredients** | Lack of particularly equipment for producing a good yield of Soy milk and fine textured Soy flour. Labor intensity of methods<br>Lack of ingredients particularly for making confectionary products and Soy sausage.<br>Lack of time to prepare soybean products, which may depend on household size and later result into preparation neglect | Soybean processing |
| **Soybean value chain governance** | Low emphasis on soybeans in the food system. Poor Soybean value chain governance | Soybean policy and regulation |
| **Gender** | Low male involvement | Soybean Processing |
| **Enabling Environment and Food Environment** |  |  |
| **Availability of Soybean for household consumption** | Due to poor access to inputs like seed, inoculum, fertilizer | Soybean Production |
|  | Lack of access to affordable credit services | Soybean production |
| **Accessibility of Soybean for household consumption** | Accessibility of whole soybeans for consumption throughout the year was negligible due to lack of information, low male involvement, and lack of land, poor soils and expensive seed. | Soybean production |
| **Safety and Health** | No heat treatment of soybean in some households producing a product with possible anti-nutritional factors and beany flavor particularly Soy porridge and milk | Soybean processing |
|  | Use of refined mealie affects protein balance | Soybean utilization |
| **Convenience** | Hard to cook characteristics especially when cooked whole. | Soybean processing and utilization |
| **General Environmental Factors** |  |  |
| **Land and soil** | Lack of land in some wards | Production and utilization |
|  | Poor soils in some wards |  |

### Recommendations and Priorities

Immediately, the following strategies could be explored. Intensifying Soybean growing not only for sale but also for home consumption in order to enhance availability, accessibility and ultimately processing and utilization. Wards with poor markets, poor soils, and lack of land could be market avenues for locally processed soybean products. The wards could also be encouraged to grow small portions for consumption. Re-examining male involvement in Soybean Processing and Utilization. Low levels of male involvement may be responsible for tendencies of selling the entire soybean grown without leaving any for family meals. In this study, male headed households grew more soybean but accessibility to whole soybean for processing and utilization was negligible in both male headed and female headed households. Female-headed households as well as smaller sized households should also be supported to grow soybean as a cash crop as well as for family consumption. Strengthening educating farmers on the benefits of soybeans as well as how to make various soybean products. This could encourage them to grow it not only as a cash crop, but also for household consumption. Dangers of anti-nutritional factors and how to destroy them in soybeans to ensure protein digestibility should also be among the key messages. Standardizing soybean household processes could further help the households to be correctly processing soybean. This could also lead to correct knowledge diffusion in the non-GIZ areas thereby leading to producing quality products in all the wards. Plant protein complementarity and use of whole maize meal in diverse diets should equally be promoted particularly through cooking demonstrations [40]. Market linkages of farmers to seed companies need to be created and strengthened in order for them to access inputs at fair price. Farmers themselves should also become more involved in soybean seed production and multiplication. Encouraging farmers who cannot afford certified seed to use recycled seed. Use of inoculum and compound D were reported to be helpful in poor soils. In addition, more companies should be encouraged to venture into soybean value addition leading to market led growth and utilization at household level. Extension services for soybean processing and utilization should be improved. This could be achieved through training the camp officers, rural health facility outreach staff as well as community volunteers correct processing of soybean. They could also be educated on how to make various soybean products. Use of household and privately owned equipment in the community should be encouraged as a sustainable approach to soybean utilization. Utilizing appropriate technologies in the community at a fee such as hammer mills should be encouraged. Existing harmer mill operators could be oriented on how to mill fine Soy flour by repeat milling. In this study soybean flour produced using local harmer-mills was reported to be of a coarse texture. Lastly there is need to advocate among stake holders for more emphasis on soybeans in the food system Promoting other rural enterprises that will have equipment to process whole soybeans at a fee such as soybean extruders to produce soy pieces in close vicinity could equally bring about market led growth and utilization in the medium to long run. Soy pieces were consumed by almost every household in Eastern Province. Other equipment that could be promoted include; Soybean oil expeller or press and Soybean blenders or pulverizing equipment for making Soy milk.

## Data Availability

All relevant data are within the manuscript and its Supporting Information files.

## Acknowledgements

We would like to thank the Government of the Republic of Zambia through the Ministry of Agriculture for their direct support to this study through the District Agricultural Coordinators and the Camp officers. Many thanks also go to the Ministry of Health particularly Rural Health Facilities for releasing some staff to be part of the enumerators. Special thanks go to Survey enumerators from five Wards in Petauke namely Chilimanyama, Msumbazi, Nyika, Mateyo Mzeka and Ongolwe. Others are from five wards in Katete specifically; Mkaika, Katiula, Mwandafisi, Chimutende and Matunga. There were also five camp officers from Chitando, Makangila, Msanga, Nthope and Mboza Wards in Chipata. Our appreciation go to the headmen who participated in Focus group discussions (FGDs). These were from Chilimanyama, Musumbazi, Katiula, Chimutende, Nthope and Makangila wards. We should equally give credit to the two Engineers; Schultz Shangala and Funduluka Shangala the assistant data managers and analysts for formulating the structured questionnaire using the KoboCollect v2022.2.3 and later analyzing the data. Lastly, we would like to thank the Levy Mwanawasa Medical University for the support in this noble course.

